# Clinical Course and Outcomes of coronavirus disease 2019 (COVID-19) in Rheumatic Disease Patients on Immunosuppression: A case Cohort Study at a Single Center with a Significantly Diverse Population

**DOI:** 10.1101/2020.10.26.20219154

**Authors:** Timothy Arleo, David Tong, Julie Shabto, Ghazala O’Keefe, Arezou Khosroshahi

## Abstract

**Objectives:** To determine clinical course and outcomes in rheumatic disease patients with coronavirus disease 2019 (COVID-19) and compare results to uninfected patients.

**Methods:** We conducted a case cohort study of autoimmune disease patients with COVID-19 (confirmed by severe acute respiratory syndrome coronavirus 2 PCR) from 02/01/2020 to 07/31/2020 and compared them in a 1:3 ratio with uninfected patients who were matched based on race, age, sex, and comorbidity index. Patient demographics, clinical course, and outcomes were compared among these patient groups.

**Results:** A total of 70 rheumatic disease patients with COVID-19 (mean age, 56.6 years; 64% African American) were identified. The 34 (49%) patients who were hospitalized used oral glucocorticoids more frequently (p<0.01). All 10 patients on anti-TNFα medications were treated as outpatients (p<0.01). Those hospitalized with COVID-19 more often required ICU admission (17 (50%) vs 27 (26%), OR=2.78 (95% CI: 1.24 to 6.20)) and intubation (10 (29%) vs 6 (6%), OR=6.67 (95% CI: 2.20 to 20.16)) than uninfected patients. They also had higher mortality rates (6 (18%) vs 3 (3%), OR=7.21 (95% CI: 1.70 to 30.69)). Of the six COVID-19 patients who died, one was of African ancestry (p=0.03).

**Conclusions:** Rheumatic disease patients infected with COVID-19 were more likely to require ICU admission, ventilation, and died more frequently versus uninfected patients with autoimmune disease. Patients on anti-TNFα medications were hospitalized less frequently while those on chronic glucocorticoids were hospitalized more frequently. These findings have important implications for medication choice in rheumatic disease patients during the ongoing spread of COVID-19.

## INTRODUCTION

Data on COVID-19 in patients with immune-mediated rheumatic diseases using immunosuppressive medications such as biologic and conventional disease-modifying anti-rheumatic drugs (DMARDs), chronic glucocorticoids, and others is limited. Recent literature suggests that DMARDs may provide protective effects^1-2^ as they have been hypothesized to disrupt the cytokine storm reaction that often leads to serious disease^3^. Some mixed data exists regarding chronic glucocorticoid use and COVID-19 infection, with recent large database analysis showing increased rates of hospitalization due to compromised immunity^4-5^. We aim to leverage our large and diverse population of rheumatic disease patients with high prevalence of African American patients infected with COVID-19 to further characterize outcomes based on immunosuppressive medication.

Patients with rheumatic disease and those on immunosuppressive medications are considered at higher risk for contracting severe COVID-19 disease, and several groups have studied outcomes in COVID-19 patients with rheumatic disease^4,6-7^. In hospitalized rheumatic disease patients, one potential confounder is that rheumatic disease patients admitted to the hospital experience poor outcomes regardless of COVID-19 infection^8-9^. Studies have reported that up to 1/3 of admitted rheumatic disease patients require intensive care unit (ICU) admission, and 2/3 of the ICU patients require mechanical ventilation^10-11^. We hope to further characterize the impact of COVID-19 infection on this patient population by comparing infected patients to uninfected rheumatic disease patients who are admitted to the hospital for other reasons.

## METHODS

### Study population

We conducted case cohort study on rheumatic disease patients with a positive PCR test result for COVID-19 through the Emory Healthcare System. We used a data warehouse search to identify patients who were diagnosed with an autoimmune disease, were on chronic immunosuppressants, and were seen at Emory between 02/01/20 to 7/31/20. Search criteria for autoimmune disease was based on a standardized list of autoimmune diseases (supplemental table 1). Autoimmune disease diagnoses were determined by the electronic health record (EHR) documentation and confirmed by study authors. Immunosuppressive medications considered can be found in supplemental table 2. Patients were considered immunosuppressed if, prior to a positive PCR test or hospitalization, they had taken their daily medications as prescribed for ≥ 1 month. Patients receiving injections or infusions must have received their last dose according to their prescribed medication schedule.

**Table 1:** Baseline characteristics and manifestations in rheumatic disease patients with COVID-19 who were hospitalized (n=34) versus treated outpatient (n=36).

| **Characteristic** | **All COVID-19**<br>**(n=70)** | **Inpatient**<br>**COVID-19 (n=34)** | **Outpatient**<br>**COVID-19 (n=36)** | **P value** |
| --- | --- | --- | --- | --- |
| Female n (%) | 56 (80%) | 27 (79%) | 29 (81%) | 1.00 |
| Age in years, mean<br>[95% CI] | 56.6<br>[52.4 - 60.7] | 65.2<br>[59.1 - 71.2] | 48.4<br>[44.1 - 52.7] | 0.00 |
| Race, n (%) |  |  |  | 0.94* |
| Caucasian | 20 (29%) | 9 (26%) | 11 (31%) |  |
| Black or African<br>American | 45 (64%) | 22 (65%) | 23 (64%) |  |
| Other** | 5 (7%) | 3 (9%) | 2 (6%) |  |
| **Comorbidities, n (%)** |  |  |  |  |
| Pulmonary disease*** | 11 (16%) | 8 (24%) | 3 (8%) | 0.11 |
| Diabetes | 16 (23%) | 10 (29%) | 6 (17%) | 0.26 |
| Renal disease | 14 (20%) | 12 (35%) | 2 (6%) | 0.00 |
| Cancer | 7 (10%) | 2 (6%) | 5 (14%) | 0.43 |
| Hypertension | 43 (61%) | 24 (71%) | 19 (53%) | 0.22 |
| Coronary artery disease | 7 (10%) | 6 (18%) | 1 (3%) | 0.22 |
| Congestive heart failure | 11 (16%) | 9 (26%) | 2 (6%) | 0.02 |
| Obesity, BMI 30+ kg/m2 | 28 (40%) | 13 (38%) | 15 (42%) | 0.81 |
| Smoking | 21 (30%) | 10 (29%) | 11 (31%) | 1.00 |
| Former | 20 (29%) | 10 (29%) | 10 (28%) |  |
| Current | 1 (1%) | 0 (29%) | 1 (3%) |  |
| **AI diagnosis, n (%)** |  |  |  |  |
| RA | 26 (37%) | 13 (38%) | 13 (36%) | 1.00 |
| SLE | 8 (11%) | 5 (15%) | 3 (8%) | 0.47 |
| PMR or GCA | 7 (10%) | 7 (21%) | 0 (0%) | 0.00 |
| IBD | 6 (9%) | 0 (0%) | 6 (17%) | 0.03 |
| Sarcoidosis | 5 (7%) | 2 (6%) | 3 (8%) | 1.00 |
| Vasculitis | 3 (4%) | 2 (6%) | 1 (3%) | 0.61 |
| ILD | 2 (3%) | 2 (6%) | 0 (0%) | 0.23 |
| Sjogren's syndrome | 2 (3%) | 1 (3%) | 1 (3%) | 1.00 |
| Mixed SLE and MCTD | 2 (3%) | 0 (0%) | 2 (6%) | 1.00 |
| Castleman<br>disease/TAFRO | 1 (1%) | 1 (3%) | 0 (0%) | 1.00 |
| MCTD | 1 (1%) | 1 (3%) | 0 (0%) | 0.49 |
| Scleroderma | 1 (1%) | 0 (0%) | 1 (3%) | 1.00 |
| Psoriatic arthritis | 1 (1%) | 0 (0%) | 1 (3%) | 1.00 |
| Adult onset still's<br>disease | 1 (1%) | 0 (0%) | 1 (3%) | 1.00 |
| Autoimmune hepatitis | 1 (1%) | 0 (0%) | 1 (3%) | 1.00 |
| Antisynthetase<br>syndrome | 1 (1%) | 0 (0%) | 1 (3%) | 1.00 |
| Spondyloarthropathy | 1 (1%) | 0 (0%) | 1 (3%) | 1.00 |
| Ankylosing spondylitis | 1 (1%) | 0 (0%) | 1 (3%) | 1.00 |
| **Medications, n (%)** |  |  |  |  |
| Oral glucocorticoid | 37 (53%) | 24 (71%) | 13 (36%) | 0.00 |
| >10mg/day | 21 (30%) | 14 (41%) | 7 (19%) | 0.07 |
| Hydroxychloroquine | 20 (29%) | 10 (29%) | 10 (28%) | 1.00 |
| Biological DMARDs | 15 (21%) | 3 (9%) | 12 (33%) | 0.02 |
| anti-TNFα | 10 (14%) | 0 (0%) | 10 (28%) | 0.00 |
| Rituximab | 5 (7%) | 3 (9%) | 2 (6%) | 0.67 |
| Conventional synthetic DMARDs | 27 (39%) | 12 (35%) | 15 (42%) | 0.63 |
| Methotrexate | 12 (17%) | 5 (15%) | 7 (19%) | 0.75 |
| Mycophenolate mofetil | 5 (7%) | 4 (12%) | 1 (3%) | 0.19 |
| Leflunomide | 2 (3%) | 2 (6%) | 0 (0%) | 0.23 |
| Azathioprine | 8 (11%) | 1 (3%) | 7 (19%) | 0.06 |
| Targeted synthetic DMARDs | 1 (1%) | 0 (0%) | 1 (3%) | 1.00 |
| Tofacitinib | 1 (1%) | 0 (0%) | 1 (3%) | 1.00 |
| Cyclophosphamide | 1 (1%) | 1 (3%) | 0 (0%) | 0.49 |
| Sirolimus | 1 (1%) | 1 (3%) | 0 (0%) | 0.49 |
| **Presenting symptoms, n (%)** |  |  |  |  |
| Fever | 44 (63%) | 25 (74%) | 19 (53%) | 0.02 |
| Cough | 51 (73%) | 24 (71%) | 27 (75%) | 0.78 |
| Dyspnea | 26 (37%) | 18 (53%) | 8 (22%) | 0.01 |
| Myalgias | 27 (39%) | 10 (29%) | 17 (47%) | 0.23 |
| Diarrhea | 17 (24%) | 9 (26%) | 8 (22%) | 0.59 |
| Loss of taste or smell | 19 (27%) | 5 (15%) | 14 (39%) | 0.07 |

\*P value for Race was computed by comparing "Caucasian" and "Other" versus "Black or African American"
\*\*Other race includes American Indian or Alaska Native, Native Hawaiian or other Pacific Islander and not reported
\*\*\*Pulmonary disease includes asthma or COPD.

**Table 2:** Baseline characteristics in rheumatic disease patients with COVID-19 (n=34) and age, sex, race, and comorbidity index matched rheumatic disease comparators without COVID-19 (n=102).

| Characteristic | Inpatient COVID-19 (n=34) | Inpatient comparators (n=102) | P value |
| --- | --- | --- | --- |
| Female sex, n (%) | 27 (79%) | 81 (79%) | 1.00 |
| Age in years, mean [95% CI] | 65.2 [59.1 - 71.2] | 62.9 [61.3 - 64.6] | 0.52 |
| Race, n (%) |  |  | 0.92* |
| Caucasian | 9 (26%) | 31 (30%) |  |
| Black or African-American | 22 (65%) | 67 (66%) |  |
| Other** | 3 (9%) | 4 (4%) |  |
| **Comorbidities, n (%)** |  |  |  |
| Elixhauser comorbidity index, mean [95% CI] | 10.2 [6.8 - 13.5] | 9.5 [8.6 - 10.3] | 0.72 |
| Pulmonary disease*** | 8 (24%) | 23 (23%) | 1.00 |
| Diabetes | 10 (29%) | 30 (29%) | 1.00 |
| Renal disease | 12 (35%) | 38 (37%) | 1.00 |
| Cancer | 2 (6%) | 13 (13%) | 0.36 |
| Hypertension | 24 (71%) | 76 (75%) | 0.66 |
| Coronary artery disease | 6 (18%) | 17 (17%) | 0.66 |
| Congestive heart failure | 9 (26%) | 34 (33%) | 0.53 |
| Obesity, BMI 30+ kg/m2 | 13 (38%) | 32 (31%) | 0.53 |
| Smoking | 10 (29%) | 38 (37%) | 0.53 |
| Former | 10 (29%) | 29 (28%) |  |
| Current | 0 (0%) | 9 (9%) |  |
| **AI diagnosis, n (%)** |  |  |  |
| RA | 13 (38%) | 38 (37%) | 1.00 |
| SLE | 5 (15%) | 30 (29%) | 0.11 |
| PMR or GCA | 7 (21%) | 7 (7%) | 0.04 |
| Sarcoidosis | 2 (6%) | 5 (5%) | 1.00 |
| ILD | 2 (6%) | 1 (1%) | 0.15 |
| Vasculitis | 2 (6%) | 1 (1%) | 0.15 |
| Sjogren's syndrome | 1 (3%) | 4 (4%) | 1.00 |
| MCTD | 1 (3%) | 2 (2%) | 1.00 |
| Anti-synthetase syndrome | 0 (0%) | 3 (3%) | 0.57 |
| Castleman disease/TAFRO | 1 (3%) | 0 (0%) | 0.25 |
| Irritable bowel disease | 0 (0%) | 1 (1%) | 1.00 |
| Inflammatory myopathy | 0 (0%) | 1 (1%) | 1.00 |
| Inflammatory arthritis | 0 (0%) | 1 (1%) | 1.00 |
| Mixed RA and SLE | 0 (0%) | 4 (4%) | 0.57 |
| SLE, RA, and Sjogren's disease | 0 (0%) | 1 (1%) | 1.00 |
| scleroderma | 0 (0%) | 1 (1%) | 1.00 |
| Takayasu arteritis | 0 (0%) | 1 (1%) | 1.00 |
| **Medications, n (%)** |  |  |  |
| Oral glucocorticoid | 24 (71%) | 59 (58%) | 0.23 |
| >10mg/day | 14 (41%) | 38 (38%) | 0.84 |
| HCQ | 10 (29%) | 52 (51%) | 0.03 |
| Biological DMARDs | 3 (9%) | 11 (11%) | 1.00 |
| Rituximab | 3 (9%) | 3 (3%) | 0.16 |
| anti-TNF | 0 (0%) | 3 (3%) | 0.57 |
| Infliximab | 0 (0%) | 2 (2%) | 1.00 |
| Tocilizumab | 0 (0%) | 2 (2%) | 1.00 |
| Ustekinumab | 0 (0%) | 1 (1%) | 1.00 |
| Conventional synthetic DMARDs | 12 (35%) | 43 (42%) | 0.55 |
| Methotrexate | 5 (15%) | 20 (20%) | 0.62 |
| MMF | 4 (12%) | 10 (10%) | 0.75 |
| Azathioprine | 1 (3%) | 6 (6%) | 0.68 |
| Leflunomide | 2 (6%) | 2 (2%) | 0.26 |
| Mesalamine/sulfasalazine | 0 (0%) | 5 (5%) | 0.33 |
| Targeted synthetic DMARDs | 0 (0%) | 1 (1%) | 1.00 |
| Tofacitinib | 0 (0%) | 1 (1%) | 1.00 |
| Cyclophosphamide | 1 (3%) | 0 (0%) | 0.25 |
| Sirolimus | 1 (3%) | 0 (0%) | 0.25 |
| Vedolizumab | 0 (0%) | 1 (1%) | 1.00 |
| Tacrolimus | 0 (0%) | 1 (1%) | 1.00 |

\*P value for Race was computed by comparing "Caucasian" and "Other" versus "Black or African American"
\*\*Other race includes American Indian or Alaska Native, Native Hawaiian or other Pacific Islander and not reported
\*\*\*Pulmonary disease includes asthma or COPD.

### COVID-19 case identification

COVID-19 infected patients were determined by selecting patients from the study population that received a positive severe acute respiratory syndrome coronavirus 2 (SARS-CoV-2) PCR test through Emory Medical Laboratories. If a patient was admitted, hospitalization was documented. Patients who were previously symptomatically infected with COVID-19 confirmed at an outside location and received a subsequent positive PCR test at Emory were excluded if the clinical course of their original infection was not documented.

### Matched control identification

For patients who were admitted and had a positive PCR test result, a set of matched control patients was generated in a 1:3 ratio. Control patients were selected from rheumatic disease patients who were hospitalized with no clinical concern for COVID-19 or who received a documented negative PCR test for COVID-19. Patients were matched on sex, age within 10 years, race, and Elixhauser comorbidity index within 10 of the COVID-19 inpatient group. Patients without a specified race were not matched based on race. For those with more than 3 matches, matches were prioritized based on Elixhauser score followed by age. If 3 matches were not found, search criteria were expanded to age within 20 years and Elixhauser score within 20. Patients were excluded from this study if they were hospitalized for elective procedures, psychiatric conditions, or if they had received transplanted organs.

Peak laboratory values were collected during the first hospitalization of admitted patients. The neutrophil-to-lymphocyte ratio (NLR) was calculated from the lymphocyte and neutrophil count.

### Data collection

Variables analyzed in this study were extracted from the EHR using manual review. For all patients, we collected data on demographics, primary rheumatic disease, other medical comorbidities, immunosuppressive regimen, symptoms at time of either hospital admission (inpatient COVID-19 and matched control group) or PCR test date (outpatient COVID-19 group), and date of death if indicated. For admitted patients, length of hospitalization, complications (including ICU admission, CRRT therapy, and ventilator use), and discharge disposition were collected.

### Statistical analysis

Continuous variables were analyzed using a two-tailed Welch t-test assuming unequal variance and are presented as mean (95% CI). Categorical variables were analyzed using a 2-tailed Fisher test and are presented as number (percentage). Odds ratios with corresponding 95% CI were also calculated for some outcomes of interest.

## RESULTS

From 02/01/20 to 7/31/20, there were a total of 6536 patients who tested positive for COVID-19. Of these, seventy patients had diagnosed rheumatic disease and were being treated with immunosuppressive medications, seen in table 1.

The most common autoimmune diagnoses were rheumatoid arteritis (26, 37%), systemic lupus erythematosus (SLE) (8, 11%), polymyalgia rheumatica (PMR) or giant cell arteritis (GCA) (7, 10%), irritable bowel disease (IBD) (6, 9%), and sarcoidosis (5, 7%). Of the COVID-19 positive patients, 45 (64%) were African American.

### Admitted COVID-19 versus outpatient, COVID-19 population

In total, 34 (49%) of the 70 COVID-19 patients were hospitalized. The distribution of sex and race was similar between the two groups (p = 1.00 and 0.77, respectively), but hospitalized patients were older than outpatients with mean ages of 65.2 versus 48.4 (p < 0.01) (table 1). African American patients were hospitalized at similar rates to the overall population, with 22 of 45 (49%) being admitted. Hospitalized patients were more likely to have comorbidities of renal disease and congestive heart failure (p < 0.01 and p = 0.02, respectively). All seven PMR or GCA patients were hospitalized while all six IBD patients were treated as outpatients (p < 0.01 and p = 0.03, respectively). Hospitalized patients were more likely to be receiving chronic glucocorticoids and were less likely to be taking a biologic DMARD (bDMARD) (p <0.01 and p = 0.02, respectively). Of the patients taking bDMARDs, none of the ten patients receiving anti-TNFα treatment were admitted (p < 0.01). Hydroxychloroquine (HCQ) and other chronic immunosuppressive medication use were equal among inpatient and outpatient infections. Admitted patients more frequently experienced fever and dyspnea (p = 0.02, p = 0.01 respectively) as presenting symptoms. No other symptoms differed in statistically significant frequencies between inpatient and outpatient infected patients.

### Admitted, COVID-19 versus admitted, uninfected patients

Comparing admitted rheumatic disease patients with COVID-19 patients to matched rheumatic disease comparators without COVID-19, the distribution of sex, age, and race was similar, as seen in table 2 (p = 1.0, p = 0.52, p = 0.78, respectively).

The frequency of comorbidities, including the Elixhauser comorbidity indices, were similar between the two groups. There was a larger proportion of PMR or GCA patients who were infected with COVID-19 (p = 0.04) but otherwise there were no significant differences in autoimmune diagnoses. Non-infected patients more frequently received HCQ (p = 0.03), otherwise medication use was similar between the patient groups. COVID-19 patients were more likely to report symptoms of fever, cough, myalgias, and loss of taste or smell versus their comparators (p < 0.01 for all). As seen in table 3, reported symptoms of dyspnea were similar between the groups (p=0.47). COVID-19 patients had higher maximum C-reactive protein (CRP) values and lower minimum albumin values during their first hospitalization (p < 0.01 and p = 0.02, respectively), but other laboratory values were similar between patient groups.

**Table 3:**
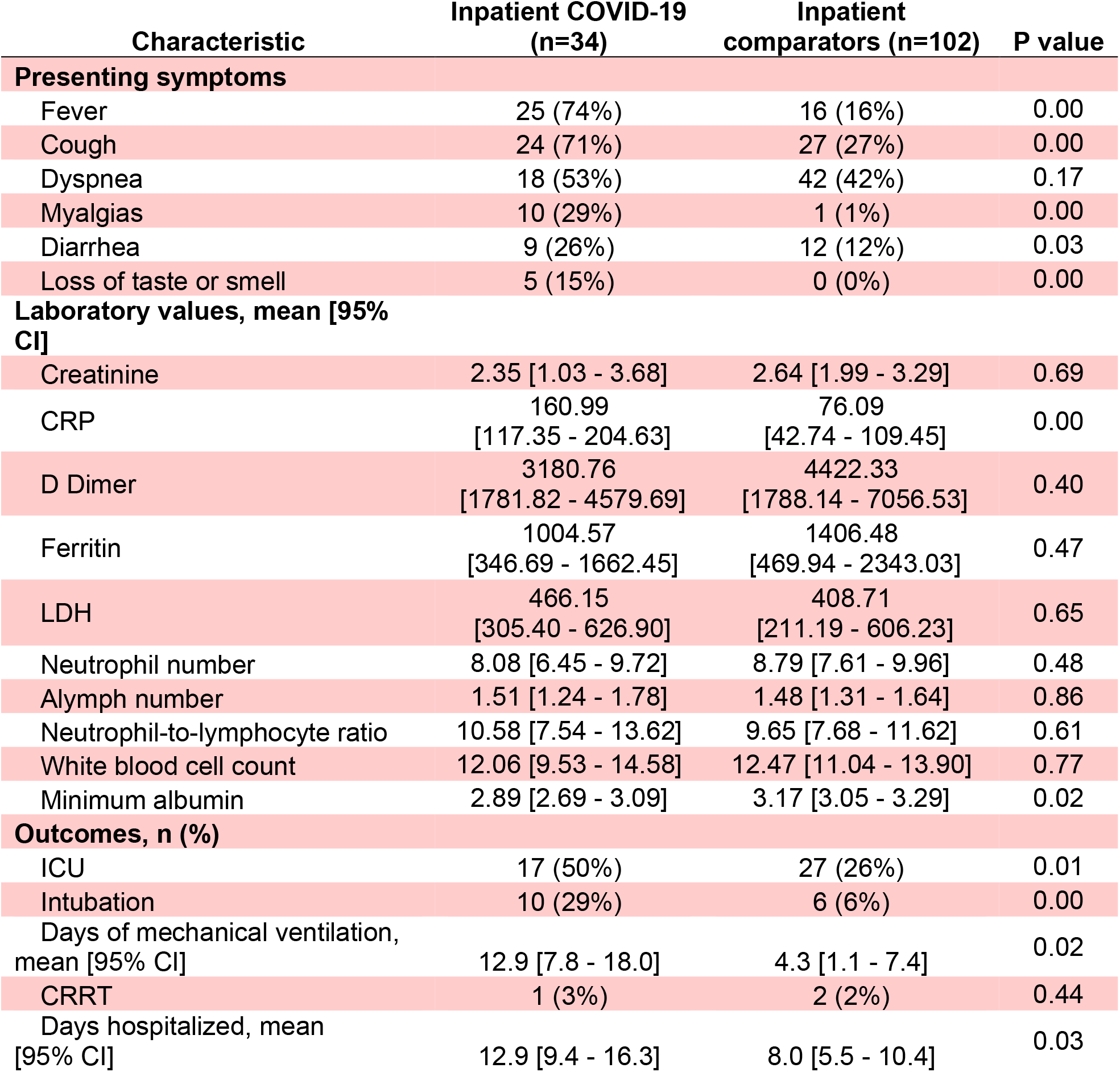
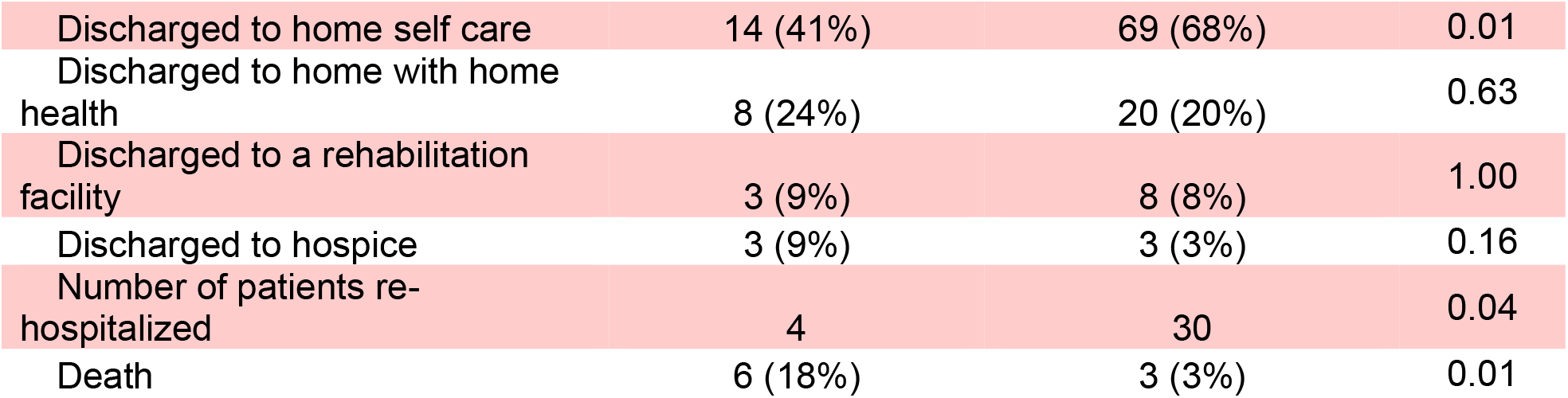
Manifestations and outcomes in hospitalized rheumatic disease patients with COVID-19 (n=34) and age, sex, race, and comorbidity index matched rheumatic disease comparators without COVID-19 (n=102).

Patients infected with COVID-19 had longer hospital stays of 12.9 days versus 8.0 days (p=0.03). A larger proportion of rheumatic disease patients with COVID-19 required ICU admission (17 (50%) vs 27 (26%), OR=2.78 (95% CI: 1.24 to 6.20), p=0.01) and required intubation (10 (29%) vs 6 (6%), OR=6.67 (95% CI: 2.20 to 20.16), p<0.01). Patients with COVID-19 remained on ventilators 12.9 days versus 4.3 days for uninfected patients (p = 0.02). COVID-19 patients were less likely to be discharged under self-care (14 (41%) vs 69 (68%), OR=0.33 (95% CI: 0.15 to 0.74), p<0.01) and more likely to die during their first hospitalization (6 (18%) vs 3 (3%), OR=7.21 (95% CI: 1.70 to 30.69), p<0.01) (table 3, table 4).

**Table 4.**
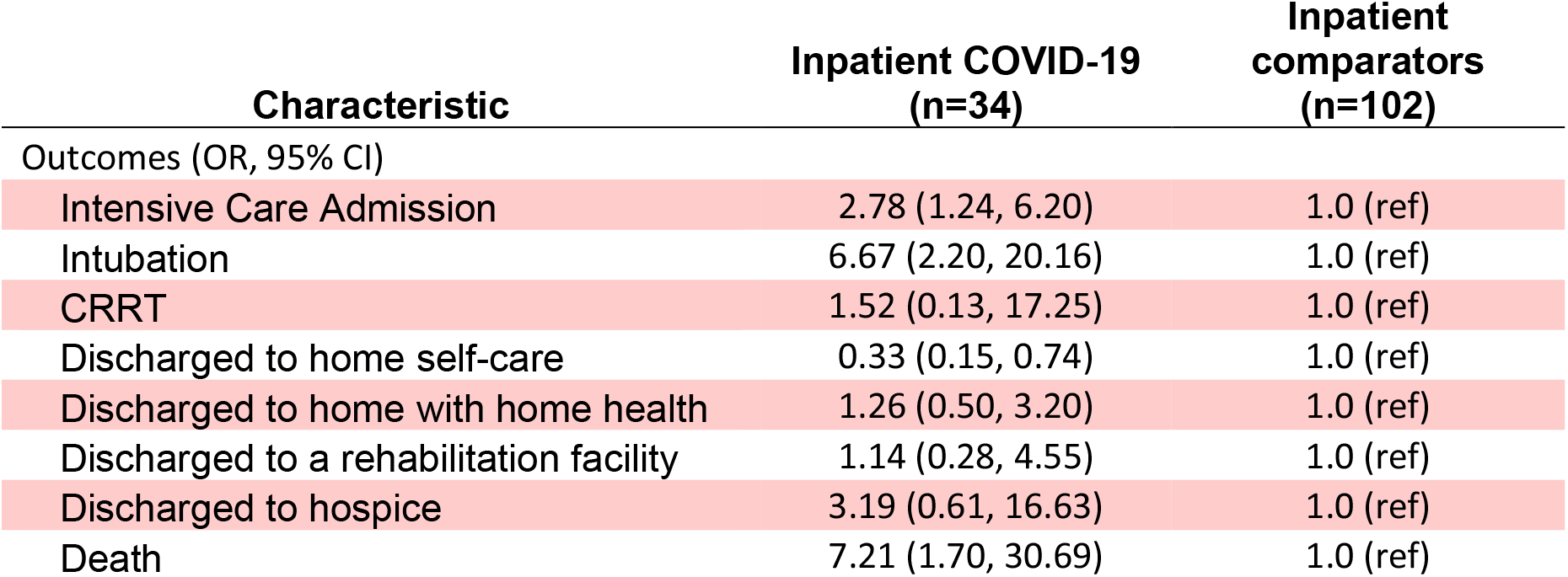
Odds ratio associations of outcomes between rheumatic disease patients with the presence and absence of COVID-19.

### Admitted, infected patients who died versus patients who survived

In total, six of the 70 COVID-19 patients died, resulting in a 9% overall mortality rate (18% mortality rate among those hospitalized). Among admitted patients with COVID-19, the distribution of sex was similar between patients who died and survived, but patients with COVID-19 who died were older with a mean age of 76.5 vs 62.8 for those that survived (p<0.01) (table 5).

**Table 5:**
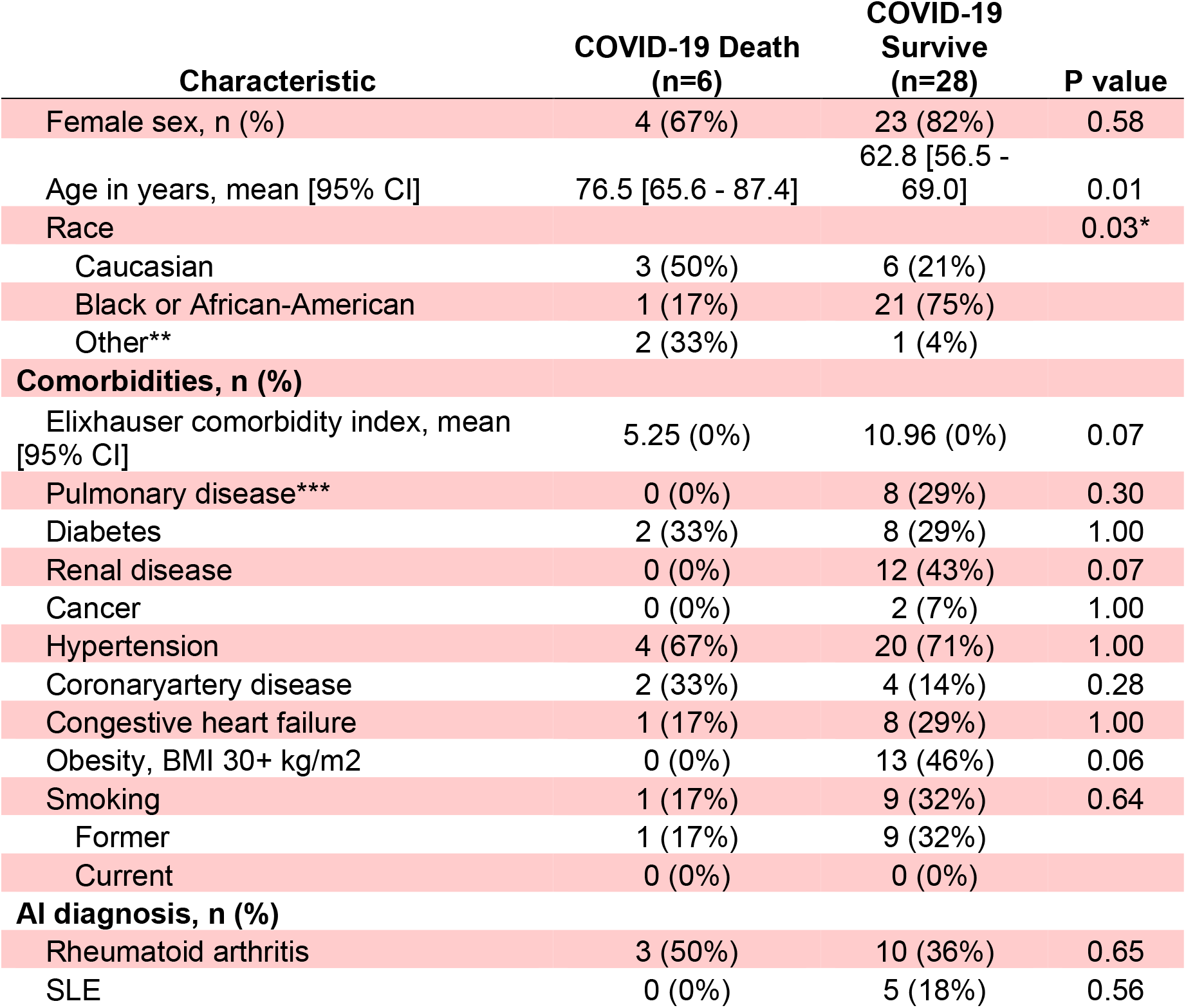
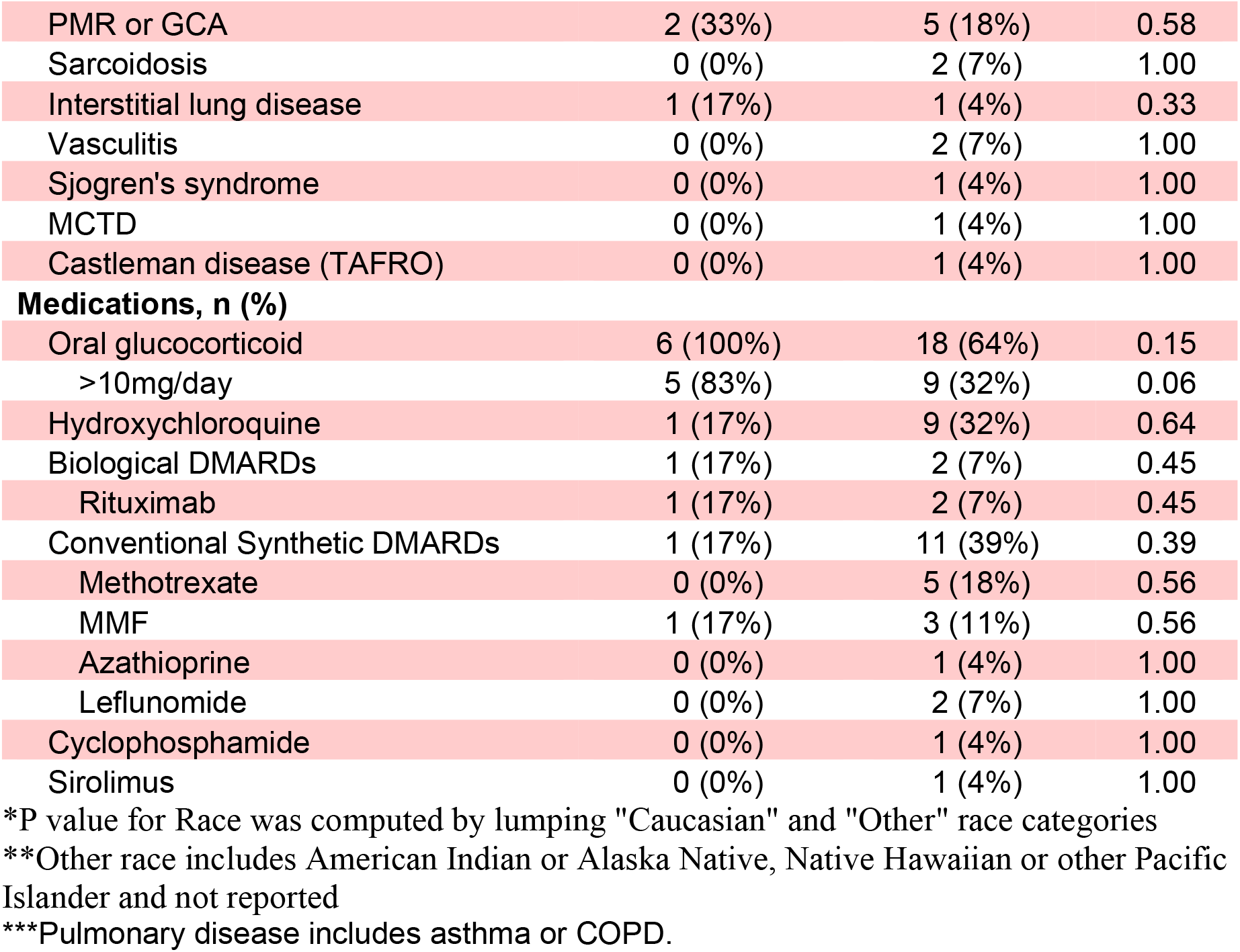
Baseline characteristics in rheumatic disease patients hospitalized with COVID-19 who died (n=6) and survived (n=28).

| Characteristic | COVID-19 Death<br>(n=6) | COVID-19<br>Survive<br>(n=28) | P value |
| --- | --- | --- | --- |
| Female sex, n (%) | 4 (67%) | 23 (82%) | 0.58 |
| Age in years, mean [95% CI] | 76.5 [65.6 - 87.4] | 62.8 [56.5 - 69.0] | 0.01 |
| Race |  |  | 0.03* |
| Caucasian | 3 (50%) | 6 (21%) |  |
| Black or African-American | 1 (17%) | 21 (75%) |  |
| Other** | 2 (33%) | 1 (4%) |  |
| **Comorbidities, n (%)** |  |  |  |
| Elixhauser comorbidity index, mean [95% CI] | 5.25 (0%) | 10.96 (0%) | 0.07 |
| Pulmonary disease*** | 0 (0%) | 8 (29%) | 0.30 |
| Diabetes | 2 (33%) | 8 (29%) | 1.00 |
| Renal disease | 0 (0%) | 12 (43%) | 0.07 |
| Cancer | 0 (0%) | 2 (7%) | 1.00 |
| Hypertension | 4 (67%) | 20 (71%) | 1.00 |
| Coronary artery disease | 2 (33%) | 4 (14%) | 0.28 |
| Congestive heart failure | 1 (17%) | 8 (29%) | 1.00 |
| Obesity, BMI 30+ kg/m<sup>2</sup> | 0 (0%) | 13 (46%) | 0.06 |
| Smoking | 1 (17%) | 9 (32%) | 0.64 |
| Former | 1 (17%) | 9 (32%) |  |
| Current | 0 (0%) | 0 (0%) |  |
| **AI diagnosis, n (%)** |  |  |  |
| Rheumatoid arthritis | 3 (50%) | 10 (36%) | 0.65 |
| SLE | 0 (0%) | 5 (18%) | 0.56 |
| PMR or GCA | 2 (33%) | 5 (18%) | 0.58 |
| Sarcoidosis | 0 (0%) | 2 (7%) | 1.00 |
| Interstitial lung disease | 1 (17%) | 1 (4%) | 0.33 |
| Vasculitis | 0 (0%) | 2 (7%) | 1.00 |
| Sjogren's syndrome | 0 (0%) | 1 (4%) | 1.00 |
| MCTD | 0 (0%) | 1 (4%) | 1.00 |
| Castleman disease (TAFRO) | 0 (0%) | 1 (4%) | 1.00 |
| **Medications, n (%)** |  |  |  |
| Oral glucocorticoid | 6 (100%) | 18 (64%) | 0.15 |
| >10mg/day | 5 (83%) | 9 (32%) | 0.06 |
| Hydroxychloroquine | 1 (17%) | 9 (32%) | 0.64 |
| Biological DMARDs | 1 (17%) | 2 (7%) | 0.45 |
| Rituximab | 1 (17%) | 2 (7%) | 0.45 |
| Conventional Synthetic DMARDs | 1 (17%) | 11 (39%) | 0.39 |
| Methotrexate | 0 (0%) | 5 (18%) | 0.56 |
| MMF | 1 (17%) | 3 (11%) | 0.56 |
| Azathioprine | 0 (0%) | 1 (4%) | 1.00 |
| Leflunomide | 0 (0%) | 2 (7%) | 1.00 |
| Cyclophosphamide | 0 (0%) | 1 (4%) | 1.00 |
| Sirolimus | 0 (0%) | 1 (4%) | 1.00 |

\*P value for Race was computed by lumping "Caucasian" and "Other" race categories
\*\*Other race includes American Indian or Alaska Native, Native Hawaiian or other Pacific Islander and not reported
\*\*\*Pulmonary disease includes asthma or COPD.

Patients who died were less likely to be African American, as one of the 22 hospitalized African American infected patients died of COVID, while three of the nine hospitalized Caucasian and two of the three hospitalized patients of another race died of COVID-19 (p=0.03). There were no differences in comorbid medical conditions between those who died and survived. Of the six patients who died, three had rheumatoid arthritis, two had PMR or GCA, and one had interstitial lung disease secondary to connective tissue disease. The frequency of these autoimmune conditions was not statistically significantly different from patients that survived. All six of the patients who died were on chronic glucocorticoids, with one being on 5mg/day and five being on doses of 10mg/day or greater. One patient who died was taking rituximab and one other patient who died was taking mycophenolate mofetil. Overall, medication use was not statistically significantly different between surviving patients and those who died. As seen in table 6, reported symptom frequency and laboratory values were similar between those who died and survived. All six patients who died were admitted to the ICU with four requiring ventilation (p < and p = 0.03, respectively).

**Table 6:** Baseline manifestations and outcomes in rheumatic disease patients hospitalized with COVID-19 who died (n=6) and survived (n=28).

| Characteristic | COVID-19 Death<br>(n=6) | COVID-19<br>Survive (n=28) | P value |
| --- | --- | --- | --- |
| **Presenting symptoms** |  |  |  |
| Fever | 4 (67%) | 21 (75%) | 0.31 |
| Cough | 3 (50%) | 21 (75%) | 0.64 |
| Dyspnea | 2 (33%) | 16 (57%) | 1.00 |
| Myalgias | 2 (33%) | 8 (29%) | 0.36 |
| Diarrhea | 0 (0%) | 9 (32%) | 0.64 |
| Loss of taste or smell | 0 (0%) | 5 (18%) | 1.00 |
| **Laboratory values** |  |  |  |
| Creatinine | 1.64 [0.78 - 2.50] | 2.50 [0.02 - 3.25] | 0.32 |
| CRP | 215.45<br>[79.72 - 351.18] | 147.38<br>[168.01 - 262.89] | 0.28 |
| D Dimer | 3402.17<br>[-2262.28 -<br>9066.61] | 3123.00<br>[1995.84 -<br>4808.49] | 0.91 |
| Ferritin | 1612.60<br>[-947.52 -<br>4172.72] | 814.56<br>[962.23 -<br>2262.97] | 0.46 |
| LDH | 456.50<br>[282.78 - 630.22] | 468.90<br>[249.01 - 663.99] | 0.92 |
| neutrophil number | 9.91 [4.02 - 15.80] | 7.74 [8.13 - 11.69] | 0.39 |
| alymph number | 1.21 [0.60 - 1.81] | 1.56 [0.90 - 1.52] | 0.22 |
| NLR | 9.26 [4.54 - 13.98] | 10.82 [5.67 -<br>12.85] | 0.53 |
| WBC count | 19.60 [9.88 -<br>29.32] | 10.44 [8.22 -<br>12.66] | 0.07 |
| minimum albumin | 2.77 [2.13 - 3.41] | 2.91 [2.54 - 2.99] | 0.60 |
| **Outcomes, n (%)** |  |  |  |
| ICU | 6 (100%) | 11 (39%) | 0.00 |
| Intubation | 4 (67%) | 6 (21%) | 0.03 |
| Days of mechanical<br>ventilation, mean (95% CI) | 15.0 [7.5 - 22.5] | 11.2 [7.6 - 14.8] | 0.39 |

| CRRT | 0 (0%) | 1 (4%) | 1.00 |
| --- | --- | --- | --- |
| Days hospitalized, mean [95% CI] | 18.7 [10.0 - 27.4] | 11.6 [15.0 - 22.4] | 0.19 |

## DISCUSSION

In this study, we evaluated patients with rheumatic conditions on chronic immunosuppressive medications who developed COVID-19 infections. Of all COVID-19 patients, 34 (49%) required hospitalization. This is a slightly higher frequency than other studies, as 46% of rheumatic patients were reported hospitalized by Gianfrancesco *et al* using the Global Rheumatology Alliance (GRA) registry while 44% of patients in the Boston area were hospitalized from D’Silva *et al*’s report. Rates of ICU admission are similar between our study and D’Silva (50% vs 48%, respectively).

Among patients with COVID-19, hospitalized patients were more likely to be older, have heart failure, and have renal disease. These are all known risk factors for more severe COVID-19 outcomes and are supported in other studies^4,6-7,12-13^. Gianfrancesco’s study found additional risk factors including hypertension or cardiovascular disease, lung disease, and diabetes, although this study has a significantly larger COVID-19 population (n=600) than ours. Regarding DMARD use, there was a clear difference in hospitalization for those taking anti-TNFα medications, with all ten patients treated as outpatients. Rituximab, the other bDMARD used by COVID-19 patients, was not associated with lower frequency of hospitalization with three out of five patients requiring hospital admission and one of these patients dying. Exactly half of the patients taking conventional synthetic DMARDs required hospitalization. Studies have hypothesized that TNF is involved in the pro-inflammatory activity of the cytokine storm that occurs in COVID-19 infection. This cytokine storm leads to end-organ damage resulting in poor patient outcomes^14-15^. Similar to Brito *et al*, our study supports this hypothesis that DMARDs, specifically the anti-TNFα medications, play a protective role in the clinical course of COVID-19 in rheumatic disease patients by disrupting this cytokine storm, resulting in a milder form of the infection. Another possible explanation for better outcomes in patients taking anti-TNFα medications is that patients who are on DMARDs usually have better control of their underlying disease with closer follow up with their rheumatologist^16^. Furthermore, these patients usually require less chronic steroid use which has been demonstrated as a known risk factor for more severe disease course^4^. Further studies examining DMARD use in patients may help to better quantify the protective effect of medical therapy on clinical outcomes in COVID-19 infections. Specifically, including a quantitative measure of rheumatic disease control or activity may help to distinguish between these two possible explanations for patients doing better on anti-TNFα medications.

With chronic glucocorticoid use being seen at almost twice the frequency among hospitalized COVID-19 patients versus those treated as outpatients in our study (71% vs 36%), our results add to the growing body of evidence suggesting that chronic glucocorticoid use as a risk factor for more severe course^4-5^. Furthermore, of the six COVID-19 patients who died, all received chronic glucocorticoids, with 5 using 10mg/day or more of prednisone. This reinforces that high doses of chronic steroid use prior to exposure to COVID-19 may be associated with worse outcomes^4^. We suspect that with a larger study population, risk of poor outcomes associated with chronic steroid use may be further stratified by daily dosing and duration of treatment.

Our study is unique as we compared outcomes among patients with rheumatic disease and COVID-19 infection to a matched group of patients with rheumatic disease who were admitted for other reasons. This provides an important clarification to outcomes of COVID-19 among this population, as rheumatic disease patients who are admitted to the hospital have poor outcomes regardless of an underlying illness^8-9^. By comparing infected rheumatic disease patients to these comparators, this study quantifies the degree to which a COVID-19 infection in this susceptible patient population impacts outcomes not explained by underlying disease burden. The higher odds of ICU admission, ventilation, and death among COVID-19 patients are particularly concerning, as these results confirm the suspicion that rheumatic disease patients infected with COVID-19 are particularly susceptible to a severe disease course and do not do well clinically. Our study is limited in that we did not examine specific reasons for hospitalization among uninfected rheumatic disease patients. We would imagine that there is significant variability in outcomes based on the acute problem causing each patient to be admitted to the hospital. Further studies that stratify this uninfected population by reason for admission would provide greater insight into stratifying risk of poor outcomes from COVID-19 versus other common reasons for admission.

The statistically significantly lower minimum albumin concentrations and higher maximum CRP values seen in hospitalized COVID-19 patients agree with other studies. Both high CRP and low albumin values have been associated with mortality, and high CRP values have been associated with intubation^17-19^.

One of our study’s strengths is that we compared outcomes in a large healthcare system with a significantly diverse population. The racial composition of our study population is enriched with 65% African Americans, which differs from D’Silva’s primarily Caucasian population (58% of patients) whereas the GRA registry did not report racial breakdown. We expected to see a higher frequency of hospitalization among African Americans infected with COVID-19, as reported in other studies^20-21^. However, the hospitalization rate among African Americans was no different than our study’s overall hospitalization rate. Furthermore, with only one of the six patients who died being African American, in our study this population was less likely to die from COVID-19. It is important to note that these results do not discount from the growing concern that African American populations face an excess disease burden, and other studies have reported an increased frequency of hospitalization^20-21^.

Our 9% mortality rate matched the mortality rate reported in the GRA registry^4^ but was slightly higher than the 6% mortality rate reported in Boston^6^. In addition to the differing racial composition among these studies, factors such as comorbid and rheumatic disease burden may impact the difference in the observed outcomes. Unlike D’Silva and Gianfrancesco’s studies, this study only includes patients actively taking immunosuppressive medications. This may result in a self-selection for a higher proportion of active rheumatic disease in our study. In addition, none of these studies specify the severity of comorbid conditions. Given that the populations studied share different demographic backgrounds, severity of comorbid conditions among each of these patient populations are likely to differ and confound the outcome results. More than half of our patients are from African ancestry who are known to have more severe autoimmune conditions^22-23^.

The strength of this study is that it is the first to evaluate the outcomes of rheumatic disease patients on active treatment for their conditions with immunosuppressive medication among a significantly diverse population with a large number of African American patients. It is also the first to compare infected, admitted rheumatic disease patients with their uninfected rheumatic disease counterparts.

This study’s power is limited by the sample population size that was collected, especially in comparing surviving COVID-19 patients to those who died. Furthermore, we did not account for severity of rheumatic conditions, severity of comorbidities, or dosage and duration of medication regimens aside from steroids. These factors all likely contribute to clinical course and outcomes in this patient population. This study builds on the growing body of information on risk factors and clinical course of COVID-19 in rheumatic disease patients.

## Data Availability

The data is maintained at Emory University on a secure server. We plan to destroy the identifiers at the earliest opportunity consistent with conduct of the research unless there is a health or research justification for retaining the identifiers.

## Competing Interests

None declared

## Funding and all other required statements

No funding sources for this manuscript.

## Patient consent for publication

Not required

## Ethical Approval

The study was determined to be exempt by the Emory University eIRB.

## Acknowledgments

None

## Key Messages

What is already known?

- Not much is known about COVID-19 outcomes in patients with underlying rheumatic disease especially compared to rheumatic disease patients hospitalized for other causes.

What does this study add?

- We found that rheumatic disease patients with COVID-19 had poorer outcomes including ICU admission, ventilation, and death compared to uninfected rheumatic disease patients.
- This study adds further support regarding protective effects of anti-TNFα medications in COVID-19 disease course as 0 of 10 of these patients required hospitalization.

How might this impact on clinical practice or future developments?

- Further studies are needed to determine factors resulting in different outcomes reported by various studies of COVID-19 in rheumatic disease patients.
- Rheumatic disease patients with COVID-19 infection may be at significantly higher risk of poor outcomes compared to uninfected rheumatic disease patients.

## Notes

**Conflicts of Interest:** None declared

### Competing Interest Statement

Dr. O'Keefe reports personal fees from EyePoint Pharmaceuticals, outside the submitted work.

### Funding Statement

No external funding was received.

### Author Declarations

A complete waiver of HIPAA authorization and informed consent has been granted by the IRB as the research poses minimal risk.

## Citations

1. Brito CA, Paiva JG, Pimentel FN, Guimarães RS, Moreira MR. COVID-19 in patients with rheumatological diseases treated with anti-TNF. Annals of the Rheumatic Diseases. Published online June 15, 2020. doi:10.1136/annrheumdis-2020-218171

2. Monti S, Balduzzi S, Delvino P, Bellis E, Quadrelli VS, Montecucco C. Clinical course of COVID-19 in a series of patients with chronic arthritis treated with immunosuppressive targeted therapies. Annals of the Rheumatic Diseases. 2020;79(5):667–668. doi:10.1136/annrheumdis-2020-217424

3. Li X, Xu S, Yu M, et al. Risk factors for severity and mortality in adult COVID-19 inpatients in Wuhan. J Allergy Clin Immunol. 2020;146(1):110–118. doi:10.1016/j.jaci.2020.04.006

4. Gianfrancesco M, Hyrich KL, Al-Adely S, et al. Characteristics associated with hospitalisation for COVID-19 in people with rheumatic disease: data from the COVID-19 Global Rheumatology Alliance physician-reported registry. Ann Rheum Dis. 2020;79(7):859–866. doi:10.1136/annrheumdis-2020-217871

5. Strangfeld A, Eveslage M, Schneider M, et al. Treatment benefit or survival of the fittest: what drives the time-dependent decrease in serious infection rates under TNF inhibition and what does this imply for the individual patient? Ann Rheum Dis. 2011;70:1914–20.

6. D’Silva KM, Serling-Boyd N, Wallwork R, et al. Clinical characteristics and outcomes of patients with coronavirus disease 2019 (COVID-19) and rheumatic disease: a comparative cohort study from a US ‘hot spot.’ Annals of the Rheumatic Diseases. 2020;79(9):1156–1162. doi:10.1136/annrheumdis-2020-217888

7. Zhao J, Pang R, Wu J, et al. Clinical characteristics and outcomes of patients with COVID-19 and rheumatic disease in China ‘hot spot’ versus in US ‘hot spot’: similarities and differences. Annals of the Rheumatic Diseases. Published online June 15, 2020. doi:10.1136/annrheumdis-2020-218183

8. Feng, X., Zou, Y., Pan, W., Wang, X., Wu, M., Zhang, M., … & Chen, Z. (2011). Prognostic indicators of hospitalized patients with systemic lupus erythematosus: a large retrospective multicenter study in China. The Journal of rheumatology, 38(7), 1289–1295.

9. Godeau, B., Mortier, E., Roy, P. M., Chevret, S., Bouachour, G., Schlemmer, B., … & Chastang, C. (1997). Short and longterm outcomes for patients with systemic rheumatic diseases admitted to intensive care units: a prognostic study of 181 patients. The Journal of rheumatology, 24(7), 317–1323.

10. Pourrat O, Bureau JM, Hira M, Martin-Barbaz F, Descamps JM, Robert R. [Outcome of patients with systemic rheumatic diseases admitted to intensive care units: a retrospective study of 39 cases]. Rev Med Interne. 2000;21(2):147–151. doi:10.1016/s0248-8663(00)88243-0

11. Nm J, Dr K, Kk G. Rheumatologic diseases in the intensive care unit: epidemiology, clinical approach, management, and outcome. Crit Care Clin. 2002;18(4):729–748. doi:10.1016/s0749-0704(02)00025-8

12. Zheng Z, Peng F, Xu B, et al. Risk factors of critical & mortal COVID-19 cases: A systematic literature review and meta-analysis. J Infect. 2020;81(2):e16–e25. doi:10.1016/j.jinf.2020.04.021

13. Sharmeen S, Elghawy A, Zarlasht F, Yao Q. COVID-19 in rheumatic disease patients on immunosuppressive agents. Seminars in Arthritis and Rheumatism. 2020;50(4):680–686. doi:10.1016/j.semarthrit.2020.05.010

14. Li, G., Fan, Y., Lai, Y., Han, T., Li, Z., Zhou, P., … & Zhang, Q. (2020). Coronavirus infections and immune responses. Journal of medical virology, 92(4), 424–432.

15. Tay, M. Z., Poh, C. M., Rénia, L., MacAry, P. A., & Ng, L. F. (2020). The trinity of COVID-19: immunity, inflammation and intervention. Nature Reviews Immunology, 1–12.

16. Ledingham, J., Gullick, N., Irving, K., Gorodkin, R., Aris, M., Burke, J., … & Jeffries, A. (2017). BSR and BHPR guideline for the prescription and monitoring of non-biologic disease-modifying anti-rheumatic drugs. Rheumatology, 56(6), 865–868.

17. Violi F, Cangemi R, Romiti GF, et al. Is Albumin Predictor of Mortality in COVID-19? Antioxidants & Redox Signaling. Published online June 11, 2020. doi:10.1089/ars.2020.8142

18. Herold T, Jurinovic V, Arnreich C, et al. Elevated levels of IL-6 and CRP predict the need for mechanical ventilation in COVID-19. Journal of Allergy and Clinical Immunology. 2020;146(1):128-136.e4. doi:10.1016/j.jaci.2020.05.008

19. Pourbagheri-Sigaroodi, A., Bashash, D., Fateh, F., et al. (2020). Laboratory findings in COVID-19 diagnosis and prognosis. Clinica Chimica Acta.

20. Rentsch, C. T., Kidwai-Khan, F., Tate, J. P., et al. (2020) Covid-19 by Race and Ethnicity: A National Cohort Study of 6 Million United States Veterans. medRxiv.

21. Yehia BR, Winegar A, Fogel R, et al. Association of Race With Mortality Among Patients Hospitalized With Coronavirus Disease 2019 (COVID-19) at 92 US Hospitals. JAMA Netw Open. 2020;3(8):e2018039–e2018039. doi:10.1001/jamanetworkopen.2020.18039

22. Ramos, P. S., Shedlock, A. M., & Langefeld, C. D. (2015). Genetics of autoimmune diseases: insights from population genetics. Journal of human genetics, 60(11), 657-664.

23. NIH Progress in Autoimmune Diseases Research. hin National Institute of Health Publication No. 05–514 (2005)

