## Supplemental Table 1 for "Clinical Course and Outcomes of coronavirus disease 2019 (COVID-19) in Rheumatic Disease Patients on Immunosuppression: A case Cohort Study at a Single Center with a Significantly Diverse Population"

**Supplemental Table 1.** Disease categories and their associated ICD-10 codes used to identify patients with rheumatic disease.

| Category | Disease (ICD-10 code) |
|---|---|
| Inflammatory arthritis | • Rheumatoid arthritis (M05%, M06%)<br>• Ankylosing spondylitis (M45%)<br>• Psoriatic arthritis or arthropathic psoriasis (L40.5) |
| Vasculitis | • Polyarteritis nodosa and related conditions (M30%)<br>• Other necrotizing vasculopathies (M31%)<br>• Unspecified arteritis (I77.6) |
| Other autoimmune diseases | • Sarcoidosis (D86%)<br>• Systemic lupus erythematosus (M32%, L93%)<br>• Dermatopolymyositis (M33%)<br>• Systemic sclerosis (M34%)<br>• Other systemic involvement of connective tissue (M35%)<br>• Crohn's disease (K50%)<br>• Ulcerative colitis (K51%)<br>• Pulmonary eosinophilia, not elsewhere specified (J82%)<br>• Other interstitial diseases (J84%)<br>• Panuveitis, sympathetic uveitis (H44.11, H44.13) |

Note: ICD-10 = international classification of disease 10$^{th}$ revision codes. The % sign indicates a wild character allowing capture of all instances of a given code.
