## Supplemental Table 2 for "Clinical Course and Outcomes of coronavirus disease 2019 (COVID-19) in Rheumatic Disease Patients on Immunosuppression: A case Cohort Study at a Single Center with a Significantly Diverse Population"

**Supplemental Table 2.** List of medications used to identify patients on immunosuppressive medications

| Criteria | Medication |
|---|---|
| Taking medication as prescribed for ≥ 1 month | • Anakinra<br>• Azathioprine<br>• Baricitinib<br>• Cyclophosphamide<br>• Hydroxychloroquine<br>• Leflunomide<br>• Methylprednisolone<br>• Methotrexate<br>• Mycophenolate mofetil<br>• Prednisone<br>• Tofacitinib |
| Has received last scheduled injection | • Abatacept<br>• Adalimumab<br>• Belimumab<br>• Certolizumab<br>• Etanercept<br>• Golimumab<br>• Infliximab<br>• Ixekinumab<br>• Natalizumab<br>• Rituximab<br>• Secukinumab<br>• Tocilizumab<br>• Ustekinumab |
